# The GLasses Against transmission of SARS-CoV-2 in the communitY (GLASSY) trial: A pragmatic randomized trial (study protocol)

**DOI:** 10.1101/2022.02.04.22270120

**Authors:** Atle Fretheim, Lars G. Hemkens, Arnfinn Helleve, Ingeborg Hess Elgersma, Petter Elstrøm, Oliver Kacelnik

## Abstract

**Background:** A systematic review of observational studies indicated that eye protection may be an effective measure to prevent SARS-CoV-2 infections. Randomized trials are needed to assess whether the observed associations are caused by protection of the eye or confounding factors such as other systematic differences between users and non-users of eye protection, co-interventions, or changes in COVID-19 incidence when comparisons were done over time.

**Methods:** Pragmatic, virtual, parallel group, 1:1 randomized, superiority trial. We will recruit and randomize participants via an online portal. The trial will be fully remote and virtual without any personal interaction between investigators and participants. All members of the public are eligible who confirm that they are at least 18 years of age, do not regularly wear glasses, have not contracted COVID-19 since December 15^th^ 2021, and are willing to be randomized to wear, or not wear glasses in public when close to other people, for a 2-week period. Persons who are dependent on visual aids but typically use contact lenses are eligible. The participants will be randomized (1:1) to wear glasses (sunglasses or other types of glasses) in public spaces when close to others (public transport, shopping centers etc.), or to the control group. The control group will be asked not to wear glasses in public spaces when close to others. The primary outcome is positive test for COVID-19. We aim to include about 25,000 participants to have a statistical power of 80% to detect a relative risk reduction of 25% for the primary outcome.

**Discussion:** Many have easy access to sunglasses or other glasses. Wearing glasses may provide eye protection and repurposing sunglasses for infection control could be a simple, readily available, environmentally friendly, safe, and sustainable infection prevention measure.

**Trial registration:** NCT05217797 (Clinicaltrials.gov)

## Introduction

### Background and rationale

The potential effectiveness of eye protective gear for infection control was pointed out over 100 years ago [1], but this simple intervention has received limited attention during the ongoing pandemic, except as part of Personal Protective Equipment (PPE) for health care workers.

A recent systematic review of studies estimating the impact of eye protection on transmission of SARS-CoV-2 identified 5 observational studies: 1 case-control study, 1 retrospective cohort study, and 3 before-after comparisons [2]. All were conducted among health workers in various settings. With one exception, all studies reported substantial associations between use of eye protection and reduced infection risk, with odds ratios ranging from 0.04 (95% CI 0.00 to 0.69) to 0.6 (0.44 to 0.81). The systematic review authors also identified three cross sectional studies examining the use of glasses in the community, all of which found a substantially lower proportions of people wearing glasses among COVID-19 patients compared to the corresponding proportion for the public in general. A case-control study from Norway, not included in the systematic review, also reported a substantially lower proportion of wearers of eyeglasses among the COVID-19 patients than in the control group, though the difference was reduced and not statistically significant (p>0.05) after multivariate adjustment [3].

In summary, the current best evidence indicates that eye protection may be an effective measure for reducing transmission risk, but randomized trials are needed to assess whether the observed associations are caused by protection of the eye or confounding factors such as other systematic differences between users and non-users of eye protection, co-interventions, or changes in COVID-19 incidence when comparison were done over time.

Many have easy access to sunglasses, and it requires little effort to use them in everyday life - after all, some of them are worn just for fashion. Repurposing sunglasses for infection control could be a simple, readily available, environmentally friendly, safe, and sustainable infection prevention measure. Those who regularly use contact lenses could forego using lenses for infection control reasons and wear the eyeglasses they typically have, instead. Again, this could be a very simple measure to prevent infections. We therefore plan to carry out a randomized trial in the community, to assess whether wearing glasses reduces the risk of SARS-CoV-2 infection.

## Objectives

Our objective is to assess whether wearing glasses has an impact on the risk of SARS-CoV-2 infection.

## Trial design

Pragmatic, parallel group, 1:1 randomized superiority trial.

## Methods: Participants, interventions and outcomes

### Study setting

Community. We will recruit participants via an online portal that will be distributed as widely as possible to the Norwegian public. The trial will be fully remote and virtual without any personal interaction between investigators and participants.

NOTE: A similar study is about to be implemented in Denmark. We are closely collaborating with colleagues in other countries to ensure that our protocols are as similar as possible, so facilitate synthesis of findings.

### Eligibility criteria

All members of the public can take part as long as they confirm that they

1. are at least 18 years of age
2. do not regularly wear glasses
3. own or can borrow glasses that they can use (e.g. sun-glasses)
4. have not contracted COVID-19 since December 15^th^ 2021.
5. do not have COVID-19 symptoms
6. are willing to be randomized to wear, or not wear glasses outside their home when close to others for a 2-week period.
7. provide informed consent

Persons who are dependent on visual aids but typically use contact lenses are eligible.

### Who will take informed consent?

Informed, electronically signed consent (MinID or BankID) will be collected on the same web-page where people can sign up to participate and consent to being randomized to the intervention or control group. Participants will also consent to the collection and use of information from questionnaires and national registers.

### Recruitment

We will promote the trial through available media outlets, and invite the public to learn more about the trial on a web-page hosted by the Norwegian Institute of Public Health. The page will include a link to the Nettskjema-platform, where those who wish to participate can receive more information and sign the consent form.

## Interventions

### Explanation for the choice of comparators

This study aims to determine the impact of wearing glasses, in real life circumstances. The intervention is to wear sunglasses or other types of glasses when close to other people outside their home (on public transport, in shopping centres etc.). Consequently, the control group will be encouraged not to wear glasses close to other people outside their home.

### Intervention description

The intervention group will be asked to wear sunglasses (or other types of glasses) when close to others outside their home, e.g. on public transport, in shopping centres etc.

### Criteria for discontinuing or modifying allocated interventions

None. The intervention period is only 14 days for each participant, and we do not plan to define specific criteria for discontinuing or modifying allocated interventions.

### Strategies to improve adherence to interventions

None. We do not plan to maintain any form of contact with the participants between trial enrollment and data collection, 3 days after the 2-week trial period.

### Relevant concomitant care permitted or prohibited during the trial

We will not give any advice about concomitant care or interventions.

### Outcomes

Day 1 of trial period is the first day the individual participates in the trial. The intervention group will be asked to wear glasses until day 14. Three days later (day 17), the participants will complete an online questionnaire.

The primary outcome is

- positive COVID-19 test result registered in public registry (from day 3 to day 17; MSIS database).

Secondary outcomes are

- health care use for respiratory symptoms (from day 3 to day 28; KPR and NPR databases)
- health care use for injuries (from day 1 to day 21; KPR and NPR databases)
- health care use (all causes) from day 1 to day 21 (data source: KPR and NPR databases)
- any positive COVID-19 test result (from day 1 to day 17; self report)
- respiratory symptoms from day 1 to day 17 (self report)
- health care use for respiratory symptoms (from day 1 to day 17; self report)
- health care use for injuries (from day 1 to day 17; self report)
- health care use (all causes) (from day 1 to day 17; self report)

### Participant timeline

Participants will be enrolled continuously during the winter/spring of 2022. The intervention period for each participant is 14 days.

**Table 1:**
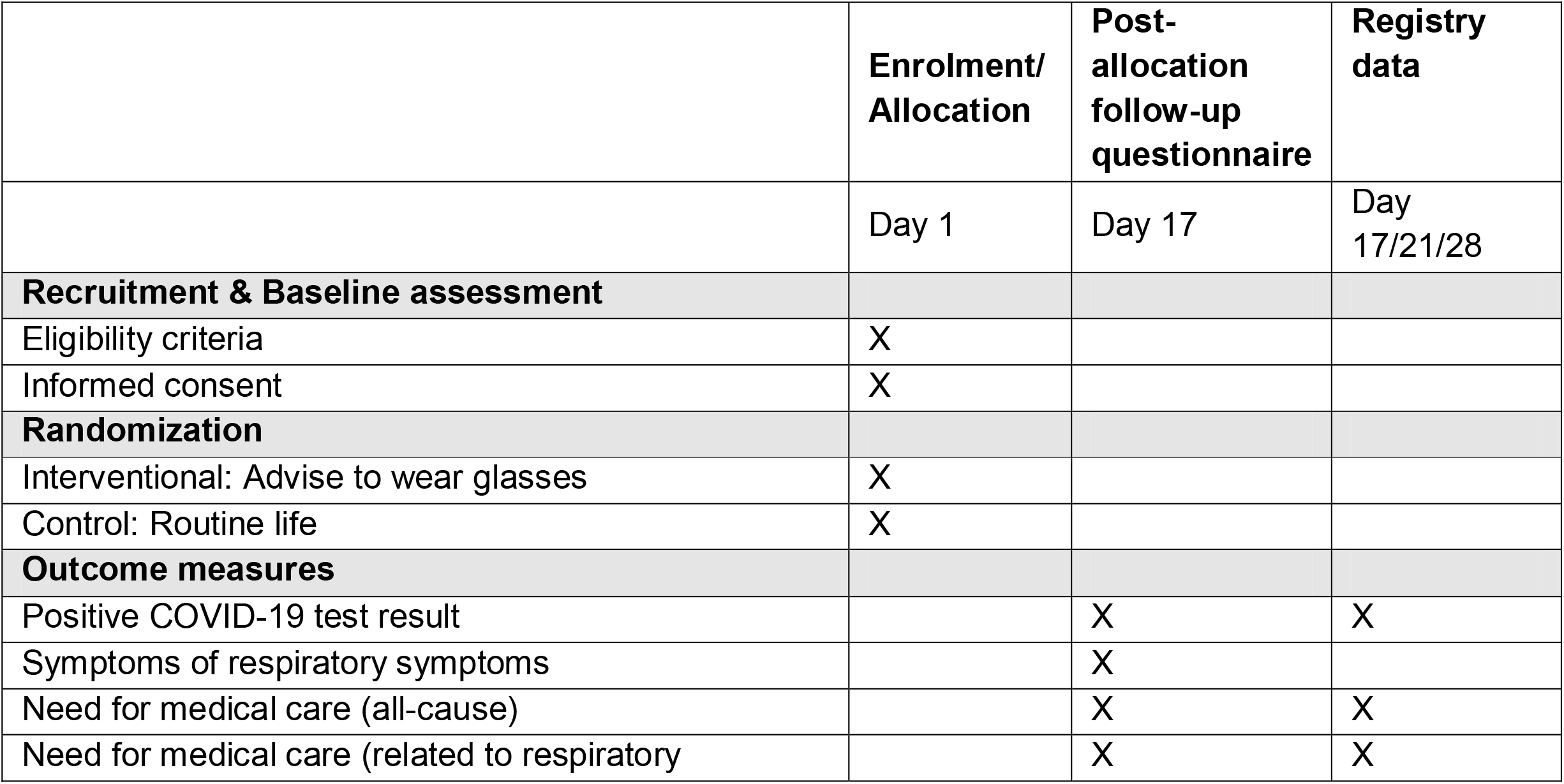
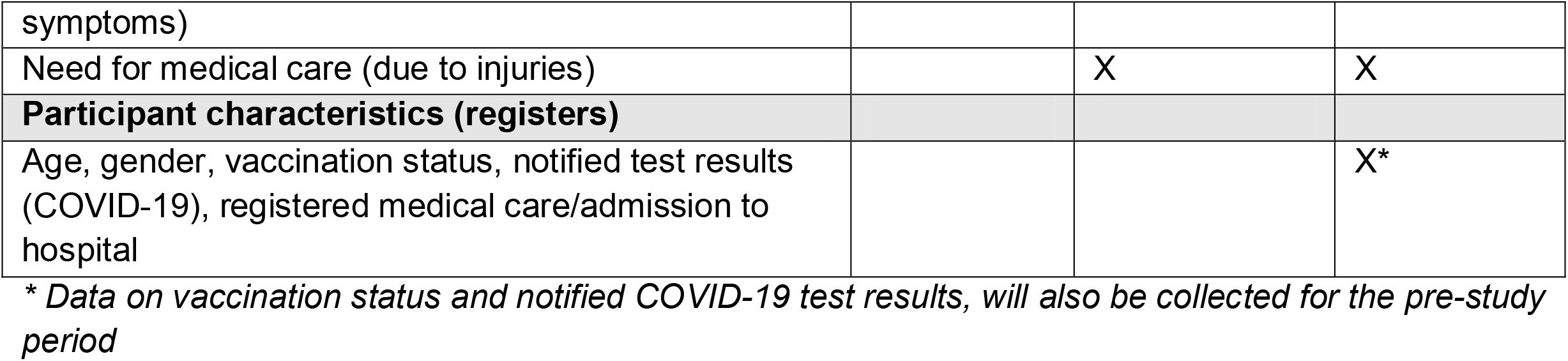
Participant timeline

### Sample size

Assuming no loss to follow-up and that wearing of glasses leads to a relative risk reduction of 25% for the primary outcome, we will need around 22,000 participants self-reporting on the primary outcome to detect a statistically significant effect (p<0.05) with 80% power. This is based on an expected event rate for the primary outcome in the control group (real life) of 2% (14-day incidence of 2000/100,000).

### Recruitment

We will recruit participants via an online portal that will be distributed as widely as possible to the Norwegian public. We will collaborate with Norwegian State Television (NRK) and other media outlets to generate public interest in the trial.

## Assignment of interventions: allocation

### Sequence generation

We will use a simple 1:1 randomization by means of computer-generated random numbers.

### Concealment mechanism

The allocation will be concealed as the participant themselves will be directly informed of their allocation as soon as they have agreed to take part in the trial and have completed the online consent form.

### Implementation

Generation of allocation sequence, enrolment of participants and assignment of participants will all be handled by the Nettskjema-platform.

## Assignment of interventions: Blinding

### Who will be blinded?

Only data analysts will be kept blind to allocation group.

## Data collection and management

### Plans for assessment and collection of outcomes

Our outcome data will be collected from national registries and from a questionnaire that the participants will be asked to complete 3 days after their trial period.

From national registries we will collect the following data:

- COVID-19 test (YES/NO) (MSIS-database)
- Result of COVID-19 test (POSITIVE/NEGATIVE) (MSIS-database)
- Health care utilization (KPR and NPR databases)
- COVID-19 vaccination status (SYSVAK).
- Age (Population registry)
- Gender (Population registry)

The questionnaire includes the following questions:

- Regular use of contact lenses?
- Use of glasses outside, in public spaces during study period? (all the time; most of the time; some of the time; never)
- Use of health care services?

### Plans to promote participant retention and complete follow-up

We will send up to two reminders to participants who have not withdrawn and who have not submitted a completed questionnaire.

### Data management

We will use the University of Oslo’s solutions for electronical signed consent form (MinID/BankID) in the web-based survey platform Nettskjema, and their secure storage of research data (TSD).

We will collect directly identifiable data (name, person identification number and e-mail address). Along with a code for linking data, the personal identification number will be sent to the registries. Each register will delete the personal identification number before register-data and code for linking are delivered and stored in TSD. The questionnaire-data are processed in the same way. The researchers linking data from all sources will not have access to the personal identification number or other direct identifiable information. The codes for linking data to the person identifier will be kept in a securely stored database, with limited access (only project leader Atle Fretheim and researcher Arnfinn Helleve will have access to this).

## Ethics

All participants will have to register and consent to participation through an online portal with information about the goal of the trial and eligibility criteria. To register they will need to use the MinID-or BankID identifier. After completing the consent form the participants will be randomized and will receive information about which study arm they have been allocated to, and they will be advised accordingly, i.e. to wear or not to wear glasses. After 17 days the participants will receive an e-mail with a link to an online data collection form.

Participation in the trial entails negligible risk and it may yield important findings that can inform decisions about infection control measures in the ongoing and future epidemics. Our assessment is that the risk-benefit of the trial is highly favorable.

The protocol has been approved by the Regional Ethics Committee of South East Norway (application number 428685).

## Statistical methods

### Statistical methods for primary and secondary outcomes

We will conduct conventional unadjusted analyses to estimate the relative risk for the prespecified outcomes, with 95% confidence intervals. The primary analysis will follow the intention-to-treat (ITT) principle (patients will be analyzed according to their group allocation).

### Methods for additional analyses (e.g. subgroup analyses)

We will conduct a subgroup analysis for participants that use contact lenses. We hypothesize that the effect may be smaller for this group, since one possible mechanism for viral transmission is that the viruses attach to ACE-2 receptors in the cornea, which we assume are protected by contact lenses (2).

We may also conduct subgroup analyses for groups with different vaccination status (number of vaccinations), and for groups that have or have not had COVID-19 previously.

### Methods in analysis to handle protocol non-adherence and any statistical methods to handle missing data

We will include all data collected, and it will be analyzed by the intention to treat principle. We do not plan to use any methods for handling missing data.

### Plans to give access to the full protocol, participant level-data and statistical code

We intend to give full access to the full protocol, participant-level dataset, and statistical code, to anyone who is interested, after securing that the dataset is fully anonymized.

## Oversight

### Adverse event reporting and harm

We will ask the participants whether they needed any medical attention during the trial period, and specifically ask for injuries.

### Plans for communicating important protocol amendments to relevant parties (e.g. trial participants, ethical committees)

We will report important protocol modifications (e.g., changes to eligibility criteria, outcomes, analyses) to the Regional Ethics Committee.

### Dissemination plans

We plan to communicate trial results to the public, and other relevant groups via publication in peer-reviewed biomedical journals, via social media, and through the Norwegian Institute of Public Health website.

## Trial status

The trial is planned to commence Feb 2^nd^, 2022.

## Data Availability

We intend to make the dataset publicly available after at has been fully anonymised.

## Declarations

### Authors’ contributions

AF is the Chief Investigator and led the protocol development. LGH had the original idea for the study and contributed to study design and to development of the protocol. AH is responsible for the data collection platform and contributed to study design and to development of the protocol. IHE, PE and OK contributed to study design and to development of the protocol.

### Availability of data and materials

The final anonymized trial dataset will be freely available to the public.

